# Defining and Assessing International Classification of Disease Suicidality Phenotypes for Genetic Studies

**DOI:** 10.1101/2024.07.27.24311110

**Authors:** Eric T. Monson, Sarah M. C. Colbert, Peter B. Barr, Cosmin A. Bejan, Ole A. Andreassen, Olatunde O. Ayinde, Zuriel Ceja, Hilary Coon, Emily DiBlasi, Anastasia Izotova, Erin A. Kaufman, Maria Koromina, Woojae Myung, John I. Nurnberger, Alessandro Serretti, Jordan W. Smoller, Murray B. Stein, Clement C. Zai, the Suicide Workgroup of the Psychiatric Genomics Consortium, Mihaela Aslan, Tim B. Bigdeli, Philip D. Harvey, Nathan A. Kimbrel, Pujan R. Patel, the Cooperative Studies Program (CSP) #572, Douglas M. Ruderfer, Anna R. Docherty, Niamh Mullins, J. John Mann

**Author notes:** Corresponding Author Contact: Eric T. Monson.

## Abstract

**Background:** Suicidality, including suicidal ideation (SI), attempt (SA), and death (SD), represents complex and partially overlapping phenotypes. This complexity contributes to study population heterogeneity in suicidality research, impeding replication efforts and data consolidation by research consortia. The standardization of suicidality definitions would help but has been insufficiently addressed in existing literature. Here, the Suicide Workgroup of the Psychiatric Genomics Consortium (PGC) provides International Classification of Disease (ICD) definitions, a critical real-world data source, for SA and SI.

**Methods:** The PGC Suicide Workgroup used published definitions coupled with expert consensus to develop ICD lists to serve as suicidality phenotype definitions. One SI and two SA lists were produced and evaluated for performance against patient screening responses in two independent cohorts (N = 9,151 and 12,621) with differing ascertainment strategies.

**Outcomes:** ICD list suicidality definitions were produced. Evaluation of generated ICD lists versus patient responses across two cohorts demonstrated varied sensitivity (15·4% to 71·1%), specificity (67·6% to 96·3%), and positive predictive values (0·57-0·92). SI ICD code performance also varied in sensitivity (29·4%-86·1%), specificity (64·2% to 90·6%), and positive predictive values (0·67 to 0·98).

**Interpretation:** Guidelines were developed to provide more consistent and comparable suicidality definitions. However, real-world application of ICD codes leads to a wide range of performance, dependent on cohort characteristics, that will need to be carefully considered in implementation. Future efforts would benefit from consistent training in use of ICD codes between sites to improve generalizability, and should include validation in diverse populations.

**Funding:** This work was funded by NIMH R01MH132733 (Mullins), R01MH132733 (Ruderfer), R01MH123619 (Docherty), R01MH123489 (Coon), R01MH124839 (PGC4), R01MH118233 and MH117599 (Smoller), Brain and Behavior Research Foundation No. 31248 (Monson), the Huntsman Mental Health Institute, National Science Foundation Graduate Research Fellowship Program Grant #1842169, and by grant # I01BX005881 and #IK6BX006523 (Kimbrel) from the Department of Veterans Affairs.

## Introduction

Suicidality represents a complex array of partially overlapping phenotypes spanning thoughts and behaviors, including suicidal ideation (SI), suicide attempt (SA), and death by suicide (SD), with differing patterns of contributors and course being reported ^1,2^. The critical need for prevention of suicide has prompted considerable and diverse research efforts. The complexity of suicidality, however, has also contributed to a wide array of definitions that hamper comparison and reproducibility across studies. Table 1 displays current international standard suicidality definitions along with aggregated phenotype names ^3,4^.

**Table 1:** Definitions of suicide and self-harm phenotypes.

| Phenotype<br>(Abbreviation) | Phenotype Definition | Aggregated Phenotype<br>(Abbreviation) |  |  |
| --- | --- | --- | --- | --- |
| Suicidal ideation (SI)<sup>a</sup> | Thoughts of engaging in suicide-related behavior<sup>3</sup>. | Suicidality/<br>Suicidal<br>thoughts<br>and<br>behaviors<br>(STBs) |  |  |
| Suicide attempt (SA)<sup>b</sup> | A non-fatal self-directed potentially injurious behavior with any intent to die as a result of the behavior. A suicide attempt may or may not result in injury<sup>3</sup>. |  | Suicidal<br>behavior<br>(SB) | Deliberate<br>self-harm<br>(DSH) |
| Suicide death (SD)<sup>b</sup> | Death caused by self-directed injurious behavior with any intent to die as a result of the behavior<sup>3</sup>. |  |  |  |
| Non-suicidal self-injury (NSSI)<sup>b</sup> | The intentional self-inflicted destruction of body tissue without suicidal intention and for purposes not socially sanctioned<sup>4</sup> |  |  |  |

<sup>a</sup> Thoughts<sup>b</sup> Behaviors

Phenotypic complexity of suicidality arises from shared and independent risk factors from genetic and environmental sources, including underlying genetic vulnerabilities, comorbid diagnoses, and lifetime exposures. SD, SA, and SI have estimated heritability in the range of 30-50% based on family and twin studies ^5,6^, though SI, compared with psychiatric disorders, may have less estimated independent heritability ^7^ than SA and SD ^8^. Like most psychiatric phenotypes, suicidality is polygenic, arising from numerous genetic variants, and requires hundreds of thousands of samples to conduct well-powered genome-wide association studies (GWAS). This sample size requires the formation of consortia, such as the Psychiatric Genomics Consortium (PGC) ^8,9^. Consortia analyses are sensitive to heterogeneity among combined samples, potentially reducing statistical power to identify meaningful and specific genetic variation associated with disease ^10^.

Consistent phenotype definitions, arising from both patient responses to instruments and real-world coding electronic health record data, stand to reduce heterogeneity and increase power. While patient responses to screening and research questionnaires provide the most ideal source for defining phenotypes for suicidality, many real-world, clinically derived samples, including those from health registries and biobanks, may not have instrument data. For these large data sources, International Classification of Disease (ICD) codes may be the only source of data from which to define phenotypes. Despite the need for consistent ICD phenotype definitions, there is very limited literature providing guidance on phenotype selection for suicidality, and no consensus yet exists for ICD codes. Implementing consistent ICD-based phenotype definitions in this field of research, as proposed here, could result in more homogeneous study populations for consortia-based efforts and substantially improve the harmonization of results across research groups. Optimally, definitions should be focused on ease of implementation within real-world data sources, including biobanks and large-scale health registries, to aid in consistency and adoption. Here, the PGC Suicide Workgroup presents a flexible set of guidelines to represent best practices in the utilization of ICD coding lists to define SA and SI phenotypes to be implemented within the field in genetic studies.

### Considerations for creating suicidality category definitions from ICD data

ICD coding options for SI are limited. Specifically, a singular code from ICD versions 9 (V62·84) and 10 (R45·851) exists, respectively. Therefore, in the absence of specific questionnaire/instrument or natural language processing data for narrative notes, these codes are the primary means of detecting SI for cohort construction.

For SA many more coding options exist. However, careful consideration must be given to potential overlap between SA and non-suicidal self-inury (NSSI) when selecting codes. This is particularly challenging when it is considered that terms like self-harm and suicidality may be used interchangeably in different settings throughout the world. Because of this complexity, determination of suicidal intent is not possible through current codes alone, and relies instead upon phenomenological characteristics of NSSI to help separate it from SA. For example, NSSI typically involves repetitive cutaneous/shallow injuries like cutting, burning, or banging the head or part of the body against something hard ^11–14^. Conversely, codes associated with SA and not NSSI, involve other methods of self-injury including poisoning (by hormones, especially insulin, antiepileptics, analgesics, psychotropics and carbon monoxide), drowning, stabbing, and more violent/lethal methods like jumping from a height or in front of a train or vehicle, hanging, and self-inflicted gunshot wounds ^15–20^. Finally, once the codes are chosen, the consistency with which they are used clinically needs to be assessed. Moreover, inclusion and exclusion of different ICD codes may allow sensitivity gains and limited specificity tradeoff when creating a definition ^21^.

## Materials and Methods

### SA and SI ICD code list construction

Only two ICD codes exist to describe SI, V62·84 (ICD-9) and R45·851 (ICD-10), and these codes were adopted as the ICD definition for SI. For SA, ICD code lists were generated by the PGC Suicide Workgroup through evaluation and modification of a previously published SA definition list from the National Center for Health Statistics (NCHS) ^22^. This list included all intentional self-harm codes from both ICD version 9 and 10. A classification system was developed by an international team of expert clinicians (Mann, Monson, Serretti, Smoller, Sokolowski, Stein) who reviewed the ICD codes from the perspective of potential for misclassification and acceptable error rate. Considerations, supported by literature discussed in the introduction to resolve intent, were also factored into code selection.

However, given the complexity of ascribing suicidal intent to any given ICD code for defining SA, two lists of classification were generated. The first ICD list was smaller and generated to serve as the primary phenotype definition for SA (SA Narrow). Codes suspected to be more likely to represent NSSI, including superficial injury by sharp objects and burning, were excluded from the SA Narrow list. ICD codes considered to be more likely to indicate SA were included in the SA Narrow list, along with codes of “undetermined” intent for injuries related to hanging, jumping from structures/high places, jumping in front of a moving vehicle, crashing a vehicle, firearm, drowning, and fire/explosives. The second ICD code list was generated to serve as a broad (SA Broad) definition of SA and NSSI for use in screening controls or as a less-specific but more sensitive definition. The SA Broad ICD list included all codes from the narrow list, as well as all intentional and undetermined self-harm ICD codes, certain accidental codes, and codes that could cover SA or NSSI, meaning they did not rule out SA.

Codes for generating ICD SA lists were obtained from separate sources for ICD-9 and ICD-10 codes, including international and United States versions of these lists. ICD-10 and ICD-10-CM (United States) code lists were obtained from https://icd.who.int/browse10/2019/en#/ and https://www.cdc.gov/nchs/icd/Comprehensive-Listing-of-ICD-10-CM-Files.htm, respectively.

ICD-9 and ICD-9-CM code lists were obtained from https://apps.who.int/iris/handle/10665/39473 and https://www.cdc.gov/nchs/icd/icd9cm.htm, respectively.

### SI and SA ICD List Validation

The codes in a total of four ICD lists, including SI, SA Narrow, SA Broad, and published SA NCHS ^22^ lists, were validated in two datasets with both ICD and Columbia Suicide Severity Rating Scale (C-SSRS) ^23^ screening, or equivalent data, available. The requirement that instrument and ICD data be collected concurrently in the same participants limited evaluation to these two datasets. Both of the validation cohorts have been previously described in detail.

Briefly, the Cooperative Studies Program #572 cohort (CSP572), entitled “Genetics of Functional Disability in Schizophrenia and Bipolar Illness Validation” was collected to evaluate genetic and other characteristics of veterans with severe psychiatric illness ^24,25^. All participants in CSP572 (N = 9,151, 86·2% male) received C-SSRS screening during the baseline diagnostic interview. In addition, individuals in the CSP572 dataset had electronic health records (EHR) that predated baseline interviewing which were also available for evaluation. Data both before (CSP572-BE) and after (CSP572-AE) enrollment were assessed to identify impact on ICD code performance, serving as proxy for “care as usual” versus systematically screened psychiatric populations. In particular, the inclusion of measures after enrollment, including after formal screening for psychiatric illnesses and suicidality history allows evaluation of the impact on performance within a more curated psychiatric sample.

The Vanderbilt University Medical Center (VUMC) cohort was obtained in one of two ways. First, through de-identified VUMC medical record data from a research-oriented data repository, Synthetic Derivative ^26^. Synthetic Derivative (VUMC-SD) represents a general clinical population seen in all care settings within the VUMC system. Records from the VUMC-SD were screened via a natural language processing (NLP) protocol to identify patients with a history of SI or SA within narrative clinical records, followed by manual validation of evidence for SI/SA (N = 1,095, 43·2% male, 56·8% female) ^27^. Second, individuals seen at a VUMC psychiatric assessment service (VUMC-Psych) who completed self-reported screening for SI (N = 11,526, 47·5% male, 52·5% female) and/or SA (N = 11,299, 47·6% male, 52·4% female) ^27^. Similar to the CSP572 cohort, these two populations provided data on ICD codes within population/general clinic obtained (VUMC-SD) and systematically screened (VUMC-Psych) populations.

All data were handled in accordance with oversight and given ethical approval from the Veterans Affairs Central Institutional Review Boards and Vanderbilt University Medical Center, as previously described.

Total counts of individuals flagged positive for each of the four SI and SA ICD lists were obtained. A positive count was given for individuals having at least one ICD code in their record from a given SI/SA list. Counts were binned into four groups using instrument screening results as the basis of comparison. Specifically, positive ICD results were binned as “true positives” if the individual had a positive instrument/screening/NLP response for prior SI/SA and “false positives” if the individual had a negative questionnaire response. Negative ICD results were similarly binned into “true” and “false” negatives based on whether the instrument data for each corresponding individual were negative or positive for prior SI or SA, respectively. These numerical count bins were then used to calculate sensitivity, specificity, and positive predictive values for each of the four evaluated ICD lists in each population using standard formulas. In addition, all results were stratified by sex.

## Results

The two proposed SA ICD lists are summarized in Tables 2 and 3 and complete lists of individual ICD codes are available in Supplemental Tables S1 and S2. For utilization of these lists it is recommended that a single occurrence of any of the listed codes in a patient’s EHR be considered a positive result. This is recommended on the basis of the relatively low prevalence of SI and SA events in free text EHR coupled with even more infrequent SI/SA ICD coding in clinical settings. In addition, the optimal use of the proposed lists for defining suicidality phenotypes should be flexible based on the composition of a given study population and aims of the study. Results in the assessed samples represent some of the possible population scenarios that may be encountered by researchers that may lead to varied performance of ICD codes.

**Table 2:**
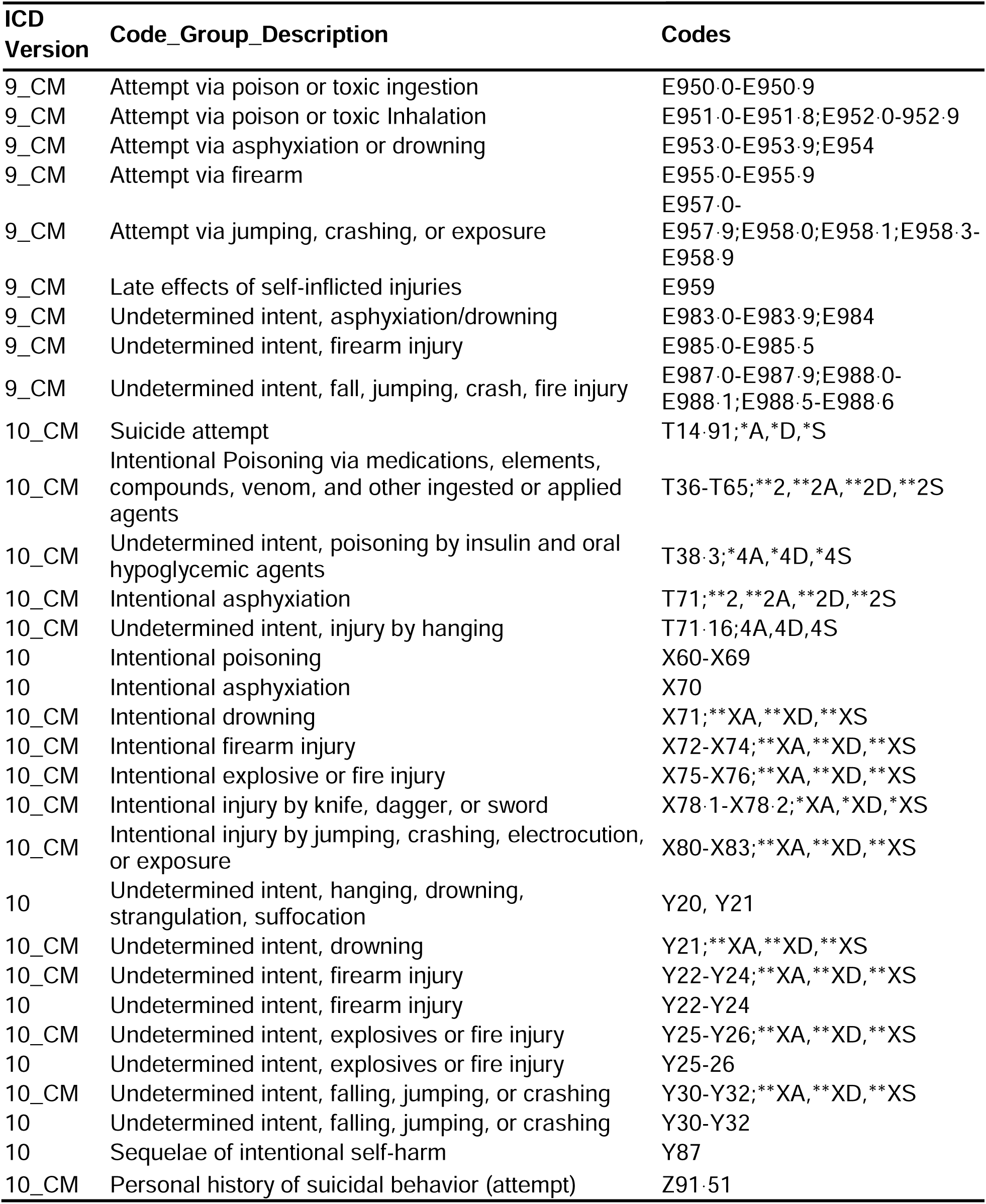
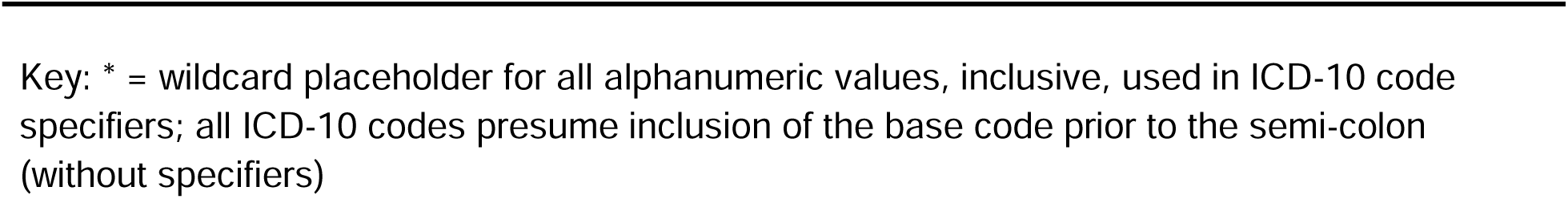
Suicide Attempt (SA) Narrow ICD Code List.

**Table 3:**
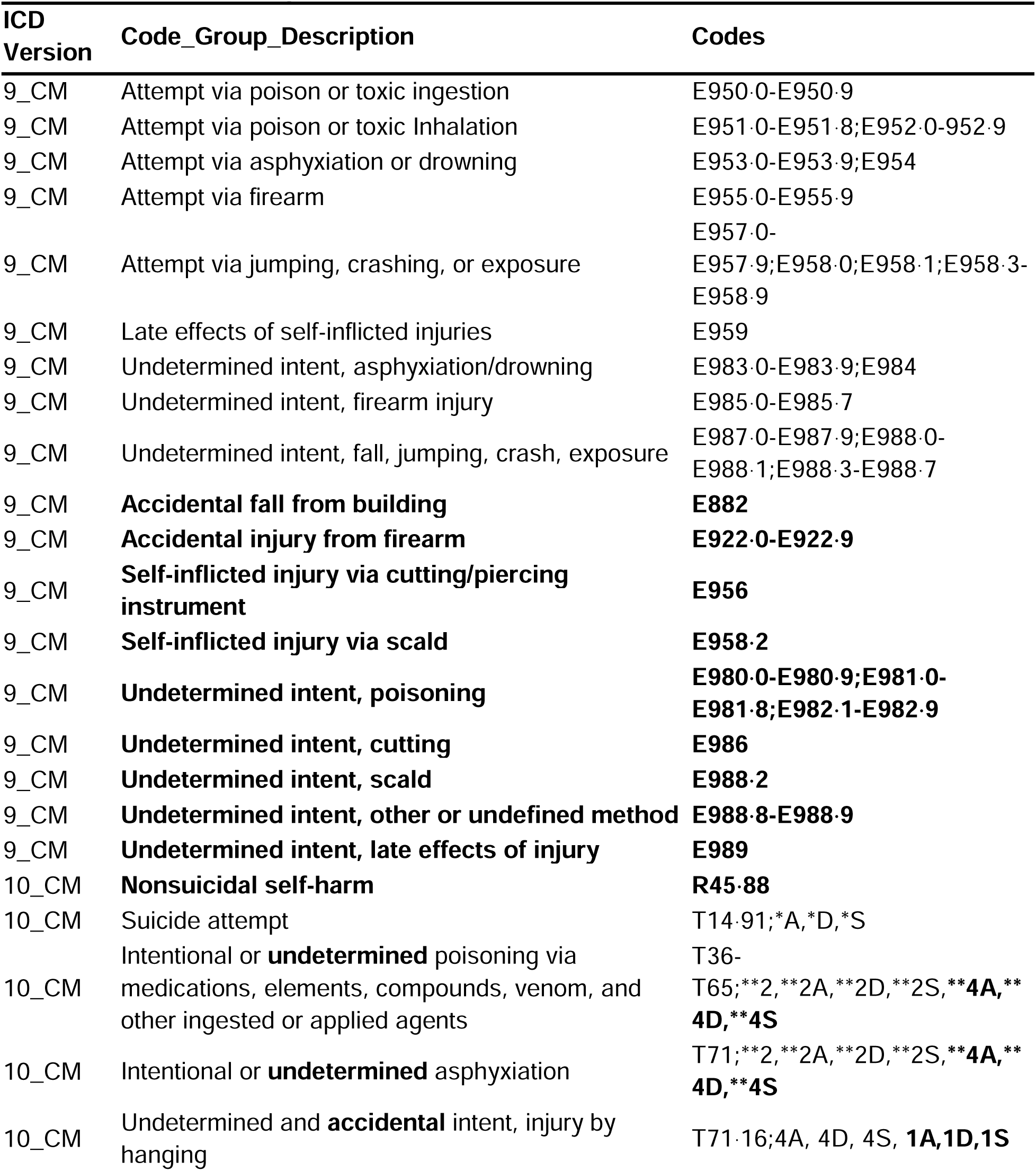
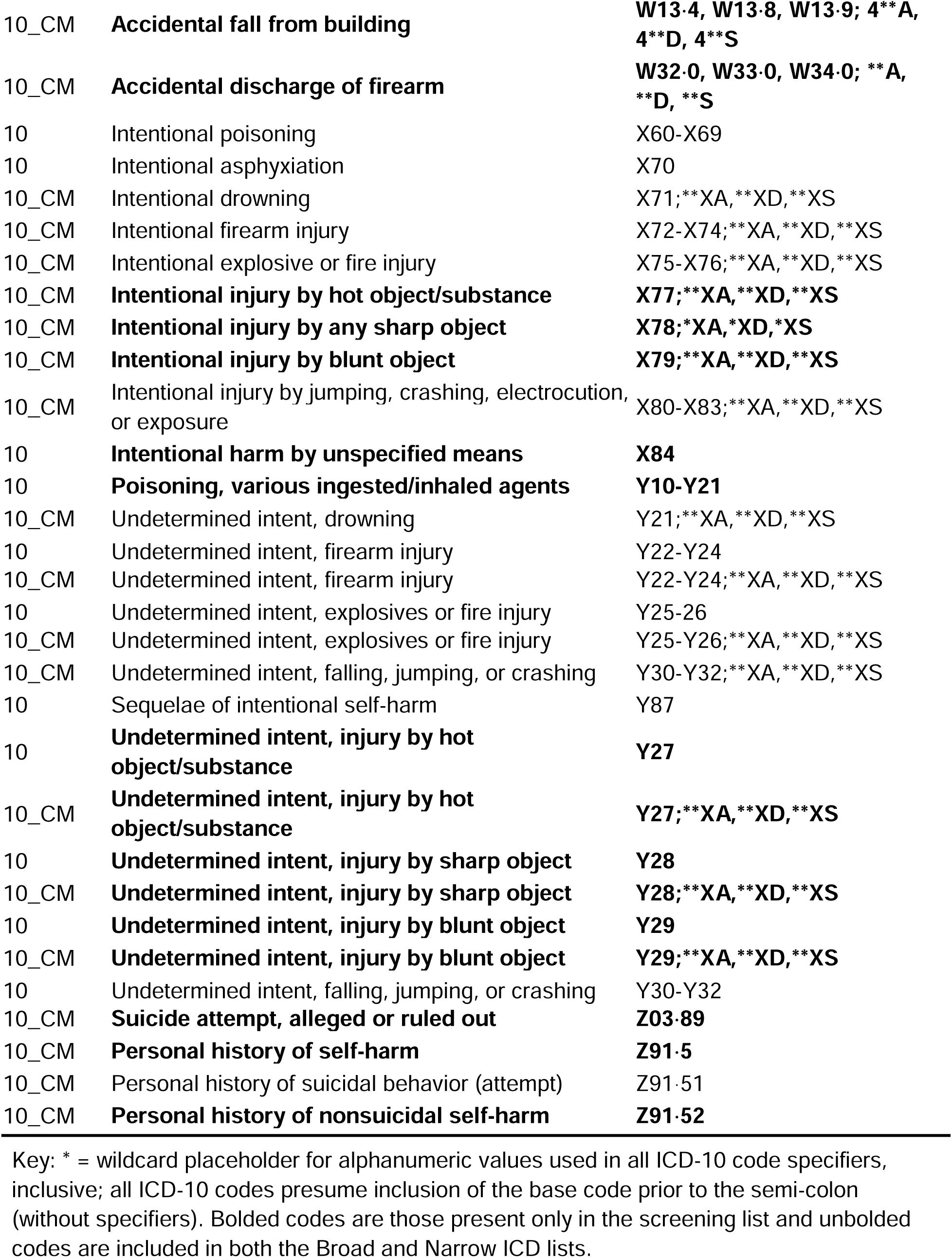
Suicide Attempt (SA) Broad ICD Code List.

The CSP572 sample represents a psychiatric population that was not selected specifically to evaluate suicidality. Such samples are common in suicidality literature, and is a likely scenario in psychiatric samples constructed from population, hospital, or national biobank efforts.

Performance within both the CSP572-BE and CSP572-AE datasets are demonstrated within Table 4. In general, the CSP572 cohorts showed low sensitivity for detection of prior events by ICD coding, with a range of 15·2%-25·6%. However, specificity was high for all ICD lists, with a range of 90·6% to 96·7%. It is notable that all three SA ICD lists showed relatively similar performance in both CSP572 samples.

**Table 4:** Suicidal Ideation and Attempt List Validation Results Versus Instrument Responses in Two Populations.

| ICD List | Cohort | TP N | FP N | TN N | FN N | SN (95% CI) | SP (95% CI) | PPV (95% CI) |
| --- | --- | --- | --- | --- | --- | --- | --- | --- |
| **SI** |  |  |  |  |  |  |  |  |
| SI Codes | CSP572-BE | 1678 | 382 | 3054 | 4037 | 29.4% (28.2%, 30.5%) | 88.9% (87.8%, 89.9%) | 0.814 (0.798, 0.831) |
| SI Codes | CSP572-AE | 2793 | 857 | 2579 | 2922 | 48.9% (47.6%, 50.2%) | 75.1% (73.6%, 76.5%) | 0.765 (0.751, 0.779) |
| SI Codes | VUMC-SD | 562 | 10 | 96 | 427 | 56.8% (53.7%, 59.9%) | 90.6% (85.0%, 96.1%) | 0.983 (0.972, 0.993) |
| SI Codes | VUMC-Psych | 4497 | 2258 | 4046 | 725 | 86.1% (85.2%, 87.1%) | 64.2% (63.0%, 65.4%) | 0.666 (0.654, 0.677) |
| **SA** |  |  |  |  |  |  |  |  |
| NCHS List | CSP572-BE | 710 | 149 | 4330 | 3962 | 15.2% (14.2%, 16.2%) | 96.7% (96.1%, 97.2%) | 0.827 (0.801, 0.852) |
| SA Narrow | CSP572-BE | 718 | 165 | 4314 | 3954 | 15.4% (14.3%, 16.4%) | 96.3% (95.8%, 96.9%) | 0.813 (0.787, 0.839) |
| SA Broad | CSP572-BE | 796 | 218 | 4261 | 3876 | 17.0% (16.0%, 18.1%) | 95.1% (94.5%, 95.8%) | 0.785 (0.760, 0.810) |
| NCHS List | CSP572-AE | 1072 | 310 | 4169 | 3600 | 23.0% (21.7%, 24.2%) | 93.1% (92.3%, 93.8%) | 0.776 (0.754, 0.798) |
| SA Narrow | CSP572-AE | 1096 | 353 | 4126 | 3576 | 23.5% (22.2%, 24.7%) | 92.1% (91.3%, 92.9%) | 0.756 (0.734, 0.778) |
| SA Broad | CSP572-AE | 1196 | 423 | 4056 | 3476 | 25.6% (24.3%, 26.9%) | 90.6% (89.7%, 91.4%) | 0.739 (0.717, 0.760) |
| NCHS List | VUMC-SD | 471 | 57 | 322 | 245 | 65.8% (62.3%, 69.3%) | 85.0% (81.4%, 88.6%) | 0.892 (0.866, 0.919) |
| SA Narrow | VUMC-SD | 415 | 36 | 343 | 301 | 58.0% (54.3%, 61.6%) | 90.5% (87.5%, 93.5%) | 0.920 (0.895, 0.945) |
| SA Broad | VUMC-SD | 509 | 123 | 256 | 207 | 71.1% (67.8%, 74.4%) | 67.6% (62.8%, 72.3%) | 0.805 (0.775, 0.836) |
| NCHS List | VUMC-Psych | 2035 | 971 | 6057 | 2236 | 47.7% (46.1%, 49.1%) | 86.2% (85.4%, 87.0%) | 0.677 (0.660, 0.694) |
| SA Narrow | VUMC-Psych | 1311 | 543 | 6485 | 2960 | 30.7% (29.3%, 32.1%) | 92.3% (91.6%, 92.9%) | 0.707 (0.686, 0.728) |
| SA Broad | VUMC-Psych | 2403 | 1836 | 5192 | 1868 | 56.3% (54.8%, 57.8%) | 73.9% (72.8%, 74.9%) | 0.567 (0.552, 0.582) |

Key: TP = true positive; FP = false positive; TN = True negative; FN = False negative; SN = Sensitivity; N = counted total number of individuals; SP = Specificity; PPV = Positive Predictive Value; SA = Suicide attempt; NCHS List = National Center for Health Statistics published ICD list; BE = ICD records collected before study enrollment; AE = ICD records obtained after study enrollment; VUMC = Vanderbilt University Medical Center; SD = Synthetic Derivative general clinical sample with identified suicidality; Psych = psychiatric urgent clinic samples; CI = confidence interval.

Comparing CSP572-BE and CSP572-AE showed differences in detection but fairly consistent performance. Specifically, records obtained after formal psychiatric screening in CSP572-AE showed increases in sensitivity as compared with CSP572-BE for SI (from 29·4% to 48·9%) and SA (15·2-17·0% to 23·0-25·6%). Positive predictive values, however, remained relatively stable for both SI (0·77-0·81) and SA (0·74-0·83).

The VUMC samples represent separate populations, VUMC-SD represents patients selected for endorsing SI/SA and seen in general clinical settings versus VUMC-Psych patients being seen in acute psychiatric settings. Overall, ICD performance in the VUMC populations was considerably different from CSP572. Sensitivity tended to be higher, ranging from 30·7% to 86·1% in VUMC SI and SA evaluation versus 15·2% to 48·9% for SI and SA evaluation in CSP572. Specificity and PPV tended to be more mixed between the samples, but were generally lower in the VUMC samples. This was especially the case for the SA screening list specificity, which had a range of 67·6-73·9% a in the VUMC samples versus 90·6-95·1% in the CSP572 samples.

Performance metrics also varied across lists used within the two VUMC populations. Overall sensitivity for SI was higher within the VUMC-Psych at 86·1% versus 56·8% in VUMC-SD. Interestingly, the opposite relationship was observed for sensitivity in SA (30·7-56·3% versus 58·0-71·1% in VUMC-Psych versus VUMC-SD). Positive predictive value, however, was consistently higher for SI and SA in VUMC-SD versus VUMC-Psych (0·567-0·677 in VUMC-Psych versus 0·805-0·983 in VUMC-SD). Of note, the proposed SA NarrowICD list was found to have higher positive predictive values and specificity than the SA Broad list, while the SA screening list produced consistently higher sensitivity values.

Sex-specific analyses of all datasets were also performed and are reported in supplemental Table S3. It is noted that, across all populations, screening for SI in males showed consistently higher sensitivity and frequently higher specificity compared with females (range of an absolute difference of 1·7-11·6% in sensitivity, and 2·1-3·3% in specificity). SI and SA positive predictive values were similar within each sample between males and females. No clear/consistent trends or differences were observed for sensitivity or specificity within SA sex-specific analyses.

## Discussion

ICD codes are an important data source that are frequently available and phenotypically relevant, and may represent the only means of classifying suicidality within a given real-world dataset. However, the utilization of ICD codes introduces many challenges that should be considered in study design and the interpretation of the results presented here. First, results presented here are consistent with other studies in finding that ICD codes are generally much less sensitive at detecting a suicidality event than instruments or NLP algorithms ^25,27,28^. In addition, ICD coding is impacted by: 1) lack of a uniform standard and scope of practice for clinician coding training; 2) variable clarity, accuracy, details enabling determination of suicidal intent, and completeness of patient history; 3) insurance billing requirements and provider policies that may influence the use of some codes for administrative purposes; 4) differences in region, institution/site, population, religion, and culture that influence stigma regarding suicidality; and 5) potential liability or legal implications of assigning a suicidality code ^29^. In addition, it is critical to note that many instances of SI and SA may not lead to formal medical evaluation and that, in some medical systems, not all codes that are applied for billing purposes are directly connected to the patient record, but could be identified if claims data are available, also leading to SA/SI that will be missed unless the patient is responding to a questionnaire.

Based on these challenges, it can be expected that the performance of ICD codes in suicidality phenotype definitions will vary considerably based on characteristics of the sample assessed. Specifically, codes captured, at best, 86% of SI cases and 71% of SA cases identified via instruments or NLP/manual review but more frequently found less than 50%. However, codes were comparatively reliable when present, and demonstrated generally good specificity and positive predictive value. Importantly, the proposed definitions performed best when samples were derived from acute psychiatric evaluation settings (VUMC-Psych) or from clinical samples enriched for suicidal phenotypes (VUMC-SD), and performed worst in more general psychiatric samples (both CSP572). However, it must be noted that ascertainment differences between samples and cultural/clinical practice differences among sites may also be substantially contributing to these observed differences. Finally, the proposed ICD definitions performed within the range of other evaluations in different populations ^30–32^, suggesting reasonably representative validation populations.

Based on these characteristics, there are multiple potential use scenarios for the presented ICD lists. For example, the SA Narrow list could be used as a highly specific but less sensitive definition to improve homogeneity of analyses, especially in larger samples that are less limited by power constraints. In such cases, the SA Broad list could be used to screen control/comparison populations. However, the SA Broad list also showed consistently higher sensitivity with only modest decrease in specificity and PPV in most of the assessed samples. As such, the Broad list could also be used as a primary phenotype definition.

Many factors, including some of the issues with ICD coding noted above, may have influenced the striking performance differences between the CSP572 and VUMC samples. For example, it has been noted that psychiatric emergency rooms routinely evaluate suicidality cases ^33^. As such, providers in these settings may be more likely to code suicidal events via ICD, leading to improved capture and ICD sensitivity, as seen in the VUMC-Psych sample. Populations routinely assessed for other, non-acute reasons, however, may be less likely to receive specific ICD coding for prior suicidal events. This may be the case within the CSP572 cohort, which represents a population assessed and managed for significant and chronic psychiatric illness. It is notable that, following instrument screening of the CSP572 cohort, the sensitivity of ICD coding did increase substantially, suggesting that incorporation of systematic screening may lead to increased suicidality ICD coding. Finally, clinical familiarity with specific populations may also change performance, such as noting differences in sensitivity and specificity in the sex-specific results. Specifically, male coding within the VA system (where the majority of patients are male) tended to have higher specificity and sensitivity in most cases. An opposite pattern, where females tended to have higher sensitivity and specificity, was observed within the VUMC populations, which had a more even male/female distribution. It is also worth noting that the evaluation of lifetime suicidality and correspond EHR codes in CSP572 predated the VA’s implementation of universal suicide risk screening in routine care settings ^34^.

Such varied results also hint at the inherent complexity of describing SI and SA, and should be considered carefully in analytical design. Specifically, separating SI and SA effectively from other forms of suicidality and NSSI is particularly difficult in the setting of ambivalence and shifting patient narrative. It is the recognition of this difficulty that prompted the adjustment of the proposed SA Narrow list to exclude events more frequently associated with NSSI, such as superficial cutaneous cutting injuries. Indeed, this likely contributed to the higher specificity seen in the use of the proposed SA Narrow in the VUMC populations. However, the overlapping nature of suicidality and NSSI phenotypes may also serve as a justification for using the SA Broad list as the basis of the primary study definition, especially when sample sizes constrain power and analytical methods.

Ultimately, the optimal strategy to define a given suicidality phenotype and control populations may vary based on available information, sample size, and the study question. Regardless of the strategy employed, a clear explanation of the rationale and design of the sample is recommended to aid future replication and meta-analysis efforts.

### Study Limitations

This study has several potential limitations. First, the proposed ICD lists are designed to be easily and widely implementable. Alternative and potentially superior strategies that make use of more complex ICD coding inclusion/exclusion algorithms, empirical assessment of codes’ association with known suicidality outcomes, natural language processing, and manual review, are not evaluated in this work. Specifically, there is evidence that the use of simple list definitions of individual ICD codes may reduce the capacity to differentiate SA and NSSI features compared with classification models that use multiple codes or EHR elements ^35^. A simple design was selected intentionally to increase portability and usability. In addition, other methods listed are not yet standardized and require more extensive access to individual records or resources than many studies may have. Future iterations of this protocol may include consideration of these strategies.

In addition, the ICD code validation was also performed only within United States samples due to limitations in existing non-US samples with the required data types, limiting generalization to international samples. Even other US samples may vary considerably due to variations in clinical documentation practices and systemic biases, including racial bias, in psychiatric diagnoses ^36^. Therefore, future work on additional large datasets that include international and more diverse ascertainment strategies would be beneficial and more generalizable.

## Conclusion

The proposed SI and SA Narrow/Broad definition ICD lists from this workgroup have been designed to improve the overall consistency of phenotyping and sample selection for suicide genetics research studies. They are intended to serve as a framework for the design of genetic and consortium SA studies, and will not be ideal for every study. ICD codes are incomplete in many cases, and supplemental information from free text EHR may greatly increase ascertainment rates of both SI and SA. However, the evaluation and discussion of the complexity of suicide-related phenotypes is a crucial consideration when establishing future samples and criteria. In addition, it is strongly recommended that any effort exploring a suicide phenotype will provide clear descriptions of selection criteria and rationale for the design on the basis of the study question with these complexities in mind. Improving the clarity and consistency of samples will be critical to identify robust and interpretable risk factors for SA and related phenotypes, in an effort to improve the identification of high-risk individuals, improve treatment modalities, reduce associated tangible and personal costs, and ultimately prevent suicide.

## Data Availability

All data produced in the present work are contained in the manuscript.

## Acknowledgements

This material is the result of work supported with resources from the Department of Veterans Affairs Cooperative Studies Program (CSP #572). The contents do not represent the views of the U.S. Department of Veterans Affairs or the United States Government.

Additional Members of the Suicide Working Group of the Psychiatric Genomics Consortium: Mark Adams, Rolf Adolfsson, Ingrid Agartz, Esben Agerbo, Tracy M Air, Martin Alda, Lars Alfredsson, Adebayo Anjorin, Vivek Appadurai, María Soler Artigas, Allison E. Ashley-Koch, Swapnil Awasthi, M Helena Azevedo, Amanda Bakian, Nicholas Bass, Claiton HD Bau, Bernhard T Baune, Jean C. Beckham, Frank Bellivier, Andrew W Bergen, Klaus Berger, Wade H Berrettini, Joanna M Biernacka, Elisabeth B Binder, Michael Boehnke, Martin Bohus, Marco P Boks, Anders D Børglum, Rosa Bosch, David L Braff, Harry Brandt, Gerome Breen, Richard Bryant, Monika Budde, Cynthia M Bulik, Enda M Byrne, Wiepke Cahn, Adrian I Campos, Miguel Casas, Enrique Castelao, Jorge A Cervilla, Xiao Chang, Boris Chaumette, Hsi-Chung Chen, Wei J Chen, Erik D Christensen, Sven Cichon, Jonathan R I Coleman, Aiden Corvin, Nicholas Craddock, David Craig, Steven Crawford, Scott Crow, Franziska Degenhardt, Ditte Demontis, Michelle Dennis, Srdjan Djurovic, Philibert Duriez, Howard J Edenberg, Alexis C Edwards, Annette Erlangsen, Tõnu Esko, Ayman H Fanous, Fernando Fernández-Aranda, Manfred M Fichter, Jerome C Foo, Andreas J Forstner, Mark Frye, Janice M Fullerton, Hanga Galfalvy, Steven Gallinger, Michael Gandal, Melanie Garrett, Justine M Gatt, Pablo V Gejman, Joel Gelernter, Ina Giegling, Stephen J Glatt, Philip Gorwood, Hans J Grabe, Melissa J Green, Eugenio H Grevet, Maria Grigoroiu-Serbanescu, Yiran Guo, Blanca Gutierrez, Jose Guzman-Parra, Jonathan D Hafferty, Lauren Hair, Hakon Hakonarson, Katherine A Halmi, Steven P Hamilton, Marian L Hamshere, Annette M Hartmann, Elizabeth R. Hauser, Michael A. Hauser, Joanna Hauser, Stefanie Heilmann-Heimbach, Akitoyo Hishimoto, Per Hoffmann, David M Hougaard, Jennifer Huffman, Hai-Gwo Hwu, Marcus Ising, Daniel Jacobson, Sonia Jain, Stéphane Jamain, Susana Jiménez-Murcia, Craig Johnson, Ian Jones, Lisa A Jones, Lina Jonsson, René S Kahn, JooEun Kang, Allan S Kaplan, Walter H Kaye, Pamela K Keel, John R Kelsoe, Kenneth S Kendler, James L Kennedy, Ronald C Kessler, Minsoo Kim, Stefan Kloiber, Kelly L Klump, Karestan C Koenen, Manolis Kogevinas, Bettina Konte, Henry R Kranzler, Marie-Odile Krebs, Po-Hsiu Kuo, Mikael Landén, Jacob Lawrence, Marion Leboyer, Phil H Lee, Daniel F Levey, Douglas F Levinson, Cathryn M Lewis, Dong Li, Qingqin S Li, Shih-Cheng Liao, Calwing Liao, Klaus Lieb, Lisa Lilenfeld, Jennifer H. Lindquist, Jolanta Lissowska, Chih-Min Liu, Adriana Lori, Susanne Lucae, Ravi Madduri, Pierre J Magistretti, Christian R Marshall, Nicholas G Martin, Fermin Mayoral, Susan L McElroy, Patrick McGrath, Peter McGuffin, Andrew M McIntosh, Benjamin McMahon, Andrew McQuillin, Sarah E Medland, Divya Mehta, Ingrid Melle, Yuri Milaneschi, James E Mitchell, Philip B Mitchell, Esther Molina, Gunnar Morken, Ole Mors, Preben Bo Mortensen, Bertram Müller-Myhsok, Richard M Myers, Caroline Nievergelt, Vishwajit Nimgaonkar, Merete Nordentoft, Markus M Nöthen, Michael C O’Donovan, Satoshi Okazaki, Catherine M Olsen, Roel A Ophoff, David W. Oslin, Ikuo Otsuka, Michael J Owen, Carlos Pato, Michele T Pato, Brenda WJH Penninx, Jonathan Pimm, Dalila Pinto, Giorgio Pistis, Renato Polimonti, David Porteous, James B Potash, Robert A Power, Abigail Powers, Martin Preisig, Xuejun Qin, Digby Quested, Josep Antoni Ramos-Quiroga, Nicolas Ramoz, Andreas Reif, Miguel E Rentería, Marta Ribasés, Vanesa Richarte, Marcella Rietschel, Stephan Ripke, Margarita Rivera, Andrea Roberts, Gloria Roberts, Stefan Roepke, Guy A Rouleau, Diego L Rovaris, Vsevolod Rozanov, Dan Rujescu, Cristina Sánchez-Mora, Alan R Sanders, Stephen W Scherer, Christian Schmahl, Peter R Schofield, Thomas G Schulze, Laura J Scott, Andrey Shabalin, Jianxin Shi, Stanley I Shyn, Lea Sirignano, Pamela Sklar, Olav B Smeland, Daniel J Smith, Marcus Sokolowski, Edmund J S Sonuga-Barke, Gianfranco Spalletta, Eli A Stahl, Anna Starnawska, John S Strauss, Fabian Streit, Michael Strober, Mei-Hsin Su, Beata Świątkowska, Laura M Thornton, Jodie Trafton, Janet Treasure, Maciej Trzaskowski, Ming T Tsuang, Gustavo Turecki, Robert J Ursano, Sandra Van der Auwera, Laura Vilar-Ribó, John B Vincent, Henry Völzke, Consuelo Walss-Bass, James TR Walters, Erin B Ware, Danuta Wasserman, Hunna J Watson, Cynthia Shannon Weickert, Thomas W Weickert, Myrna M Weissman, Frank Wendt, Thomas Werge, David C Whiteman, Leanne M Williams, Virginia Willour, Stephanie H Witt, D Blake Woodside, Naomi R Wray, Zeynep Yilmaz, Lea Zillich.

## The CSP #572 study team

Planning Committee: M. Aslan, M. Antonelli, M. de Asis, M. S. Bauer, M. Brophy, J. Concato, F. Cunningham, R. Freedman, M. Gaziano, T. Gleason, P. D. Harvey, G. Huang, J. Kelsoe, T. Kosten, T. Lehner, J. B. Lohr, S. R. Marder, P. Miller, T.J. O’Leary, T. Patterson, P. Peduzzi, R. Przygodzki, L. Siever, P. Sklar, S. Strakowski, H. Zhao.

Executive Committee: M. Brophy, J. Concato, A.H. Fanous, M. Gaziano, P.D. Harvey, T. Kosten, A. Malhotra, S. Mane, T. Bigdeli, P. Sklar, L. Siever, H. Zhao.

Study Chairs’ Offices: VA Healthcare System, Bronx, NY: L. Siever (Co-Chair), M. Corsey, L. Zaluda. VA Healthcare System, Miami, FL: P.D. Harvey (Co-Chair), J. Johnson.

CSP Epidemiology Centers: VA Clinical Epidemiology Research Center CERC, VA Connecticut Healthcare System, West Haven, CT, included J. Concato Director, Methodological Co-Principal Proponent, M. Aslan, D. Cavaliere, V. Jeanpaul, A. Maffucci, L. Mancini; the Massachusetts Veterans Epidemiology Research and Information Center MAVERIC, VA Boston Healthcare System, Jamaica Plain, MA, included M. Gaziano Director, Methodological Co-Principal Proponent, J. Deen, G. Muldoon, S. Whitbourne.

Study Sites: Albuquerque: J. Canive, L. Adamson, L. Calais, G. Fuldauer, R. Kushner, G. Toney, M. Lackey, A. Mank, N. Mahdavi, G. Villarreal. Atlanta: E. C. Muly, F. Amin, M. Dent, J. Wold. Baltimore: B. Fischer, A. Elliott, C. Felix, G. Gill. Birmingham: P. E. Parker, C. Logan, J.

McAlpine. Boston/Brockton: L.E. DeLisi,. Charleston: M.B. Hammer, D. Agbor-Tabie, W. Goodson. Cincinnati: M. Aslam, M. Grainger, Neil Richtand, Alexander Rybalsky. Houston: R. Al Jurdi, E. Boeckman, T. Natividad, D. Smith, M. Stewart, S. Torres, Z. Zhao. Indianapolis: A. Mayeda, A. Green, J. Hofstetter, S. Ngombu, M. K. Scott, A. Strasburger, J. Sumner. Little Rock: G. Paschall, J. Mucciarelli, R. Owen, S. Theus, D. Tompkins. Long Beach: S.G. Potkin, C. Reist, M. Novin, S. Khalaghizadeh. Miami: R. Douyon, J. Johnson, N. Kumar, B. Martinez.

Minneapolis: S.R. Sponheim, T.L. Bender, H.L. Lucas, A.M. Lyon, M.P. Marggraf, L.H. Sorensen, C.R. Surerus. Montrose: C. Sison, J. Amato, D.R. Johnson, N. Pagan-Howard. New York Harbor: L.A. Adler, S. Alperin, T. Leon. Northampton: K.M. Mattocks, N. Araeva, J.C. Sullivan. Palo Alto: T. Suppes, K. Bratcher, L. Drag, E.G. Fischer, L. Fujitani, S. Gill, D. Grimm, J. Hoblyn, T. Nguyen, E. Nikolaev, L. Shere, R. Relova, A. Vicencio, M. Yip. Philadelphia: I. Hurford, S. Acheampong, G. Carfagno. Pittsburgh: G.L. Haas, C. Appelt, E. Brown, B. Chakraborty, E. Kelly, G. Klima, S. Steinhauer. Salisbury: R.A. Hurley, R. Belle, D. Eknoyan, K. Johnson, J. Lamotte. San Diego: E. Granholm, K. Bradshaw, J. Holden, R. H. Jones, T. Le, I.G. Molina, M. Peyton, I. Ruiz, L. Sally. Tacoma: A. Tapp, S. Devroy, V. Jain, N. Kilzieh, L. Maus, K. Miller, H. Pope, A. Wood. Temple: E. Meyer, P. Givens, P. B. Hicks, S. Justice, K. McNair, J.L. Pena, D.F. Tharp. Tuscaloosa: L. Davis, M. Ban, L. Cheatum, P. Darr, W. Grayson, J. Munford, D. Smith, B. Whitfield, E. Wilson. Washington DC: A.H. Fanous, S.E. Melnikoff, B.L Schwartz, M.A. Tureson. West Haven: D. D’Souza, K. Forselius, M. Ranganathan, L. Rispoli.

Albuquerque, NM, CSP Coordinating Center: M. Sather Director, C. Colling, C. Haakenson, D. Krueger.

VA Office of Research and Development: T. O’Leary Chief Research and Development Officer, G. Huang Director, Cooperative Studies Program, T. Gleason Director, Clinical Science Research and Development Service, R. Przygodzki Associate Director for Genomic Medicine, and Acting Director of Biomedical Laboratory Research and Development Service, S.

Muralidhar Senior Scientific Program Manager Genomic Medicine Program, Biomedical and Clinical R&D Services.

Million Veteran Program: Consortium Acknowledgement for Manuscripts MVP Executive Committee:

- Co-Chair: J. Michael Gaziano, M.D., M.P.H.

- Co-Chair: Rachel Ramoni, D.M.D., Sc.D.

- Jim Breeling, M.D. ex-officio

- Kyong-Mi Chang, M.D.

- Grant Huang, Ph.D.

- Sumitra Muralidhar, Ph.D.

- Christopher J. O’Donnell, M.D., M.P.H.

- Philip S. Tsao, Ph.D. MVP Program Office

- Sumitra Muralidhar, Ph.D.

- Jennifer Moser, Ph.D.

MVP Recruitment/Enrollment

- Recruitment/Enrollment Director/Deputy Director, Boston

- Stacey B. Whitbourne, Ph.D.; Jessica V. Brewer, M.P.H.

- MVP Coordinating Centers

– Clinical Epidemiology Research Center CERC, West Haven – John Concato, M.D., M.P.H.

– Cooperative Studies Program Clinical Research Pharmacy Coordinating Center, Albuquerque

- Stuart Warren, J.D., Pharm D.; Dean P. Argyres, M.S.

– Genomics Coordinating Center, Palo Alto – Philip S. Tsao, Ph.D.

– Massachusetts Veterans Epidemiology Research Information Center MAVERIC, Boston

- J. Michael Gaziano, M.D., M.P.H.

– MVP Information Center, Canandaigua – Brady Stephens, M.S.

- Core Biorepository, Boston – Mary T. Brophy M.D., M.P.H.; Donald E. Humphries, Ph.D.

- MVP Informatics, Boston – Nhan Do, M.D.; Shahpoor Shayan

- Data Operations/Analytics, Boston – Xuan-Mai T. Nguyen, Ph.D. MVP Science

- Genomics - Christopher J. O’Donnell, M.D., M.P.H.; Saiju Pyarajan Ph.D.; Philip S. Tsao, Ph.D.

- Phenomics - Kelly Cho, M.P.H, Ph.D.

- Data and Computational Sciences – Saiju Pyarajan, Ph.D.

- Statistical Genetics – Elizabeth Hauser, Ph.D.; Yan Sun, Ph.D.; Hongyu Zhao, Ph.D. MVP Local Site Investigators

- Atlanta VA Medical Center Peter Wilson - Bay Pines VA Healthcare System Rachel McArdle

- Birmingham VA Medical Center Louis Dellitalia

- Cincinnati VA Medical Center John Harley

- Clement J. Zablocki VA Medical Center Jeffrey Whittle

- Durham VA Medical Center Jean Beckham

- Edith Nourse Rogers Memorial Veterans Hospital John Wells

- Edward Hines, Jr. VA Medical Center Salvador Gutierrez

- Fayetteville VA Medical Center Gretchen Gibson

- VA Health Care Upstate New York Laurence Kaminsky

- New Mexico VA Health Care System Gerardo Villareal

- VA Boston Healthcare System Scott Kinlay

- VA Western New York Healthcare System Junzhe Xu

- Ralph H. Johnson VA Medical Center Mark Hamner

- Wm. Jennings Bryan Dorn VA Medical Center Kathlyn Sue Haddock

- VA North Texas Health Care System Sujata Bhushan

- Hampton VA Medical Center Pran Iruvanti

- Hunter Holmes McGuire VA Medical Center Michael Godschalk

- Iowa City VA Health Care System Zuhair Ballas

- Jack C. Montgomery VA Medical Center Malcolm Buford

- James A. Haley Veterans’ Hospital Stephen Mastorides

- Louisville VA Medical Center Jon Klein

- Manchester VA Medical Center Nora Ratcliffe

- Miami VA Health Care System Hermes Florez

- Michael E. DeBakey VA Medical Center Alan Swann

- Minneapolis VA Health Care System Maureen Murdoch

- N. FL/S. GA Veterans Health System Peruvemba Sriram

- Northport VA Medical Center Shing Shing Yeh

- Overton Brooks VA Medical Center Ronald Washburn

- Philadelphia VA Medical Center Darshana Jhala

- Phoenix VA Health Care System Samuel Aguayo

- Portland VA Medical Center David Cohen

- Providence VA Medical Center Satish Sharma

- Richard Roudebush VA Medical Center John Callaghan

- Salem VA Medical Center Kris Ann Oursler

- San Francisco VA Health Care System Mary Whooley

- South Texas Veterans Health Care System Sunil Ahuja

- Southeast Louisiana Veterans Health Care System Amparo Gutierrez

- Southern Arizona VA Health Care System Ronald Schifman

- Sioux Falls VA Health Care System Jennifer Greco

- St. Louis VA Health Care System Michael Rauchman

- Syracuse VA Medical Center Richard Servatius

- VA Eastern Kansas Health Care System Mary Oehlert

- VA Greater Los Angeles Health Care System Agnes Wallbom

- VA Loma Linda Healthcare System Ronald Fernando

- VA Long Beach Healthcare System Timothy Morgan

- VA Maine Healthcare System Todd Stapley

- VA New York Harbor Healthcare System Scott Sherman

- VA Pacific Islands Health Care System Gwenevere Anderson

- VA Palo Alto Health Care System Philip Tsao

- VA Pittsburgh Health Care System Elif Sonel

- VA Puget Sound Health Care System Edward Boyko

- VA Salt Lake City Health Care System Laurence Meyer

- VA San Diego Healthcare System Samir Gupta

- VA Southern Nevada Healthcare System Joseph Fayad

- VA Tennessee Valley Healthcare System Adriana Hung

- Washington DC VA Medical Center Jack Lichy

- W.G. Bill Hefner VA Medical Center Robin Hurley

- White River Junction VA Medical Center Brooks Robey

- William S. Middleton Memorial Veterans Hospital Robert Striker

---

**Figure 1:**
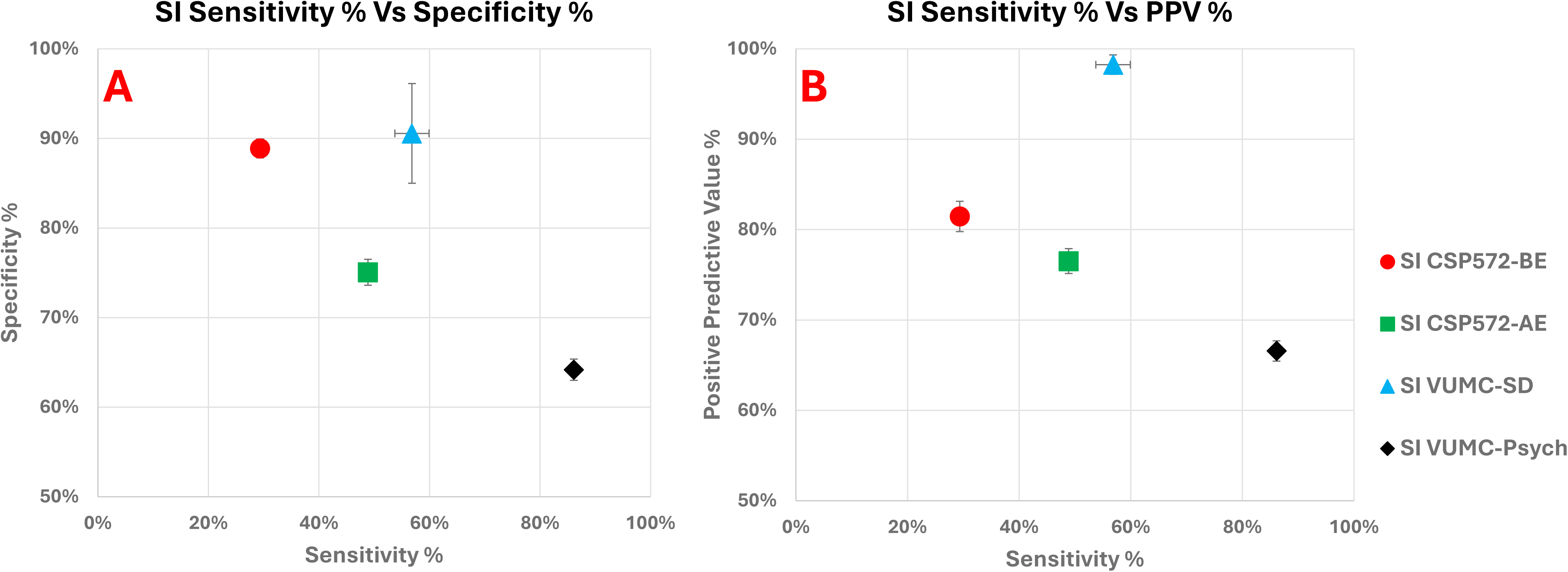
SI Sensitivity, Specificity, and PPV plots. Scatter plots representing performance metrics for suicidal ideation codes within the four evaluated samples. Panel A demonstrates plotted sensitivity vs specificity and Panel B demonstrates plotted sensitivity vs positive predictive value (PPV). 95% confidence intervals for each plotted metric are represented by whiskers. Marker colors and shapes represent the four assessed populations/timepoints, with red circles = CSP572-BE, green squares = CSP572-AE, blue triangles = VUMC-SD, and black diamonds = VUMC-Psych.

**Figure 2:**
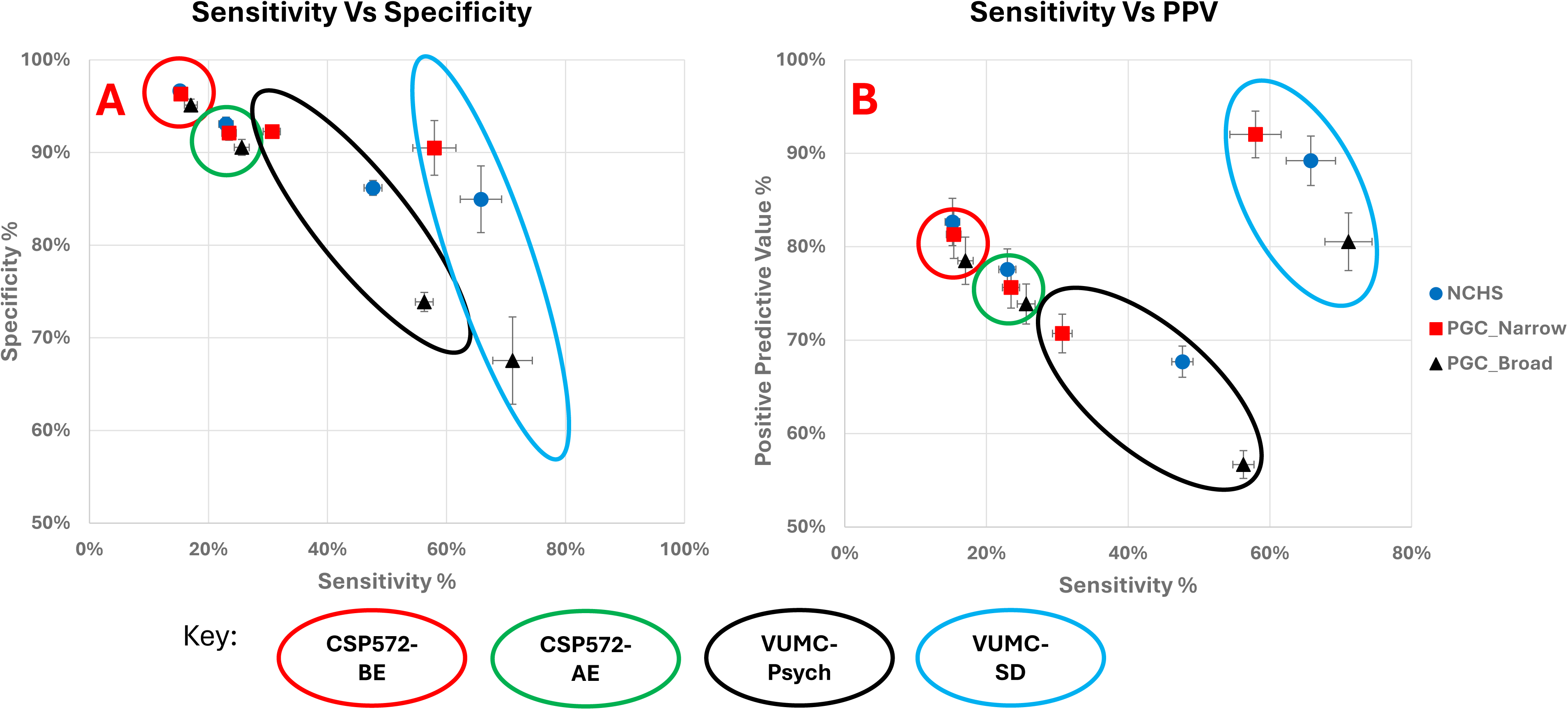
SA Sensitivity, Specificity, and PPV plots. Scatter plots representing performance metrics for suicide attempt (SA) code lists, including the published NCHS, PGC Narrow, and PGC Broad lists. Panel A demonstrates plotted sensitivity vs specificity and Panel B demonstrates plotted sensitivity vs positive predictive value (PPV). 95% confidence intervals for each plotted metric are represented by whiskers. Marker colors and shapes represent the three assessed lists, with blue circles = NCHS published list, red squares = PGC-Narrow list, and black triangle = PGC-Broad list. Colored outlines represent the four assessed populations/timepoints, with red = CSP572-BE, green = CSP572-AE, blue = VUMC-SD, and black = VUMC-Psych.

